# HIV self-testing preferences among HIV high-risk Adolescent Girls and Young Women (AGYW) in urban settings in Uganda: A Discrete Choice Experiment

**DOI:** 10.1101/2025.10.22.25338604

**Authors:** Laban Muteebwa, Patience A. Muwanguzi, Shivan Nuwasiima, Ivan Ahimbisibwe, Edson Atwine, Dan Muramuzi, Cathbert Tumusiime, Fred C. Semitala, Joanita Nangendo

## Abstract

HIV self-testing (HIVST) represents a crucial innovation for improving testing coverage, particularly among high-risk adolescent girls and young women (AGYW) inadequately reached through standard provider-assisted HIV testing approaches. Understanding user preferences for HIVST services is essential for designing models that optimize uptake. We assessed the HIVST preferences among high-risk AGYW living in urban Uganda. We enrolled high-risk AGYW consecutively in a discrete choice experiment (DCE). Each participant completed eight choice sets, each comprising of two hypothetical HIVST service alternatives and an opt-out option. The four attributes included level of assistance (assisted versus unassisted), distribution model (facility-based versus community-based), delivery channel (primary versus secondary) and sample type (oral swab versus blood). A mixed logit model was used to estimate the utility, relative importance (RI) and trade-offs. Between December 2024 and May 2025, a total of 340 participants were enrolled with mean age 19.8 years (SD: 2.6), and 65.9% of them were not in school. The distribution model, delivery channel and level of assistance significantly influenced participants’ choices to use HIVST services with coefficients of 0.14 (95%CI: 0.06, 0.23), 0.09 (95%CI: 0.00, 0.17) and 0.46 (95%CI: 0.37, 0.54) respectively. The sample type didn’t significantly influence the choice of participants to use HIVST (coefficient, 0.05 (95%CI: -0.04, 0.13). The level of assistance emerged as the most important attribute (RI: 60%), followed by facility-based distribution model (RI: 18.9%) and primary delivery channel (RI: 12.2%). AGYW were willing to sacrifice 9.2 units of utility of the test kit type to get unassisted HIVST. AGYW living in urban settings preferred unassisted HIVST delivered through the health facility. Regardless of the type of kit, the level of autonomy and convenience attained during unassisted testing greatly influenced the users’ choice of the HIVST service.

## Background

Global HIV infections have declined by 40% since 2010 – from 2.2 million to 1.3 million in 2024, yet progress remains inadequate [1]. The continued shortfall from the 2025 target of fewer than 370,000 new infections, highlighting persistent weaknesses in prevention and control efforts[1]. Women and girls continued to bear a disproportionate burden, accounting for 63% of new infections in Sub-Saharan Africa (SSA) in 2024. Within this group, adolescent girls and young women (AGYW) aged 15 – 24 years are the most severely affected: in 2024 alone, more than 4,000 AGYW were infected with HIV every week globally, with over 82.5% of these infections occurring in SSA [1]. In Uganda, the gendered impact is even more pronounced. In 2024, approximately 80% of all new HIV infections among young people aged 15 – 24 years occurred in AGYW [2]. A recent analysis of HIV testing data (November 2019 – November 2021) further revealed that, among 2,672 new infections in AGYW, at least 13.4% were recently acquired (acquired within the previous 12 months) [3]. This suggests that the majority of AGYW were living with long-standing, undiagnosed HIV infection, and therefore, not accessing timely treatment. Furthermore, one in three AGYW with recent HIV infection were first-time testers [3], underscoring the critical need to expand routine and accessible HIV testing, particularly for HIV high-risk AGYW.

HIV testing is the gateway to HIV prevention, care, and treatment, and constitutes the first pillar of the UNAIDS 95-95-95 targets: 95% of people living with HIV knowing their status, 95% of those diagnosed receiving treatment, and 95% of those on treatment achieving viral suppression [4]. In Uganda, the Ministry of Health (MoH) has expanded provider-assisted HIV testing through various approaches, including facility-based (provider-initiated and client-initiated), community outreach, home-based, and index testing [5]. Despite these efforts, conventional strategies have not optimally reached high risk populations, particularly AGYW, female sex workers, men, and men who have sex with men (MSM) [6]. Among AGYW, low HIV testing uptake are attributed to developmental factors intertwined with interpersonal, cultural, structural, and health system barriers, including stigma, limited privacy, long waiting times, negative provider attitudes, and inadequate youth-friendly services[7]. HIV self-testing (HIVST) offers a promising convenient, confidential, and youth-responsive alternative to address these challenges [8–10].

HIVST is a “test-for-triage” approach in which individuals collect their own specimen (oral fluid or blood), perform the test, and interpret the results independently or with a trusted person [11]. Evidence from randomized controlled trials and implementation studies demonstrates that HIVST significantly increases testing uptake compared with provider-delivered testing and is highly acceptable across diverse populations, including AGYW [12,13]. In Uganda, HIVST was incorporated into national HIV guidelines in 2018 [6], and is being scaled up in public and private sectors through various differentiated service delivery (DSD) models [5]. However, the heterogeneity of DSD models complicates efforts to tailor HIVST interventions to high-risk subpopulations such as AGYW and contributes to increased implementation costs [14,15]. Understanding which HIVST service delivery attributes are most valued by AGYW is therefore critical for designing AGYW-centered, cost-efficient models that optimize uptake. Prioritizing delivery models aligned with AGYW preferences has potential to enhance testing coverage and HIV positivity yield while improving resource efficiency. This study aimed to assess HIVST service preferences and key attributes associated with improved testing uptake among high-risk AGYW in urban Uganda.

## Methods

### Discrete choice experiments

This study utilized a discrete choice experiment, grounded in the Lancaster’s theory of consumer demand [16], and the Random Utility Theory (RUT) [17] that underpins modelling of choice data. Under the RUT, when an individual is presented with two or more alternatives, they will choose an alternative that maximizes utility (benefit). This latent utility is a function of; (1) a systematic (observable) component which is determined by the measurable attributes of the alternatives and the characteristics of the individual; and (2) a random (unobserved) component which captures the factors not included in the model or randomness in decision making. The utility (U_ij_) for an individual (i) choosing alternative (j) is expressed as; ***U***_***ij***_ = ***V***_***ij***_ + ***π***_***ij***_, Where; V_ij_ is the observable (deterministic) component –a linear combination of attributes and ***π***_ij_ is the unobserved (random) component. Discrete Choice Experiments (DCE) provide a structured and evidence-based method for eliciting stated preferences about health service delivery attributes [18]. By presenting participants with hypothetical service options composed of varying features (attributes), DCEs allow researchers to estimate the relative importance of each attribute, simulate uptake under different scenarios and identify trade-offs that users are willing to make [19].

### Study design and setting

This cross-sectional study was conducted among HIV high-risk AGYW living in the Kampala Metropolitan Area in Uganda from 1^st^ December 2024 to 31^st^ May 2025. This study setting was selected because prior assessment in Sub-Saharan Africa reported that adolescents and young people living in urban settings have a 1.5 to 2 times HIV prevalence compared to their counterparts in rural settings [20]. Kampala Metropolitan Area includes the four most populous districts in Central Uganda, that is, Kampala, Wakiso, Mukono and Mpigi according to the 2024 National census [21]. This is the most urbanized and commercial area in Uganda and includes its Capital City, Kampala, hosting an estimated daytime population of about five million people [22]. Moreover, this study setting accounted for the largest number of new HIV cases among AGYW of which Wakiso had the highest (4,900) followed by Kampala (2,800), Mukono (700) and Mpigi (405) [4]. Additionally, the HIV prevalence among adults aged 15-49 years (2023) in Kampala, Wakiso, Mpigi and Mukono is 7.4%, 7.2%, 8.2% and 5.3% respectively, all above the national prevalence (2023) of 5.1% [4].

### Study population

The study population included AGYW aged 15 to 24 years who had lived in the study area for at least six months, with an HIV risk score of ≥2 (high-risk), were willing to accept HIVST if it was provided (willingness score >21) and gave written informed consent (for adults and emancipated minors) or assent for minors in addition to their guardian’s consent. The HIV risk was assessed using the HIV risk assessment tool previously used among AGYW in Kampala, Uganda [24] and follows the criteria set by the MoH. The willingness to accept HIVST if it was provided was measured using the theoretical framework of acceptability [23]. This framework has seven constructs for which each construct was assessed using a one 5-level Likert item question weighted 1 – 5 for which a higher score indicated a higher level of acceptability. The sum of weights from all seven constructs was computed and a score above the 50^th^ percentile of the possible scores (21) ranging from 7 – 35 was used as a threshold of willingness to accept HIVST. Participants who couldn’t read or comprehend basic English or Luganda were excluded from the study.

### Sample size estimation and sampling procedure

The sample size was estimated using the Orme and Johnson formula [24] 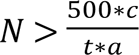 where, “N” is the estimated sample size, “c” is the number of levels per each attribute (2), “t” is the number of tasks (8) and “a” is the number of alternatives per task excluding the null alternative (opt-out) (2). Adjusting for 10% non-response and design effect of two, the estimated sample size was 140. However, this DCE study was nested in a larger study that assessed the willingness to use HIVST among HIV high-risk AGYW. All participants that demonstrated willingness to use HIVST were enrolled into the DCE study, thus a total 340 AGYW were included.

The sampling was done on three levels – firstly, two (Kampala and Wakiso) out of four districts were selected purposively because they had the highest number of new HIV infections among AGYW. Secondly, two (Makindye and Rubaga) out of five divisions in Kampala district and two (Entebbe and Nansana) out of four divisions in Wakiso district were selected using simple random sampling. Lastly, the study participants were selected using consecutive sampling technique, and the number of participants from each district was proportionate to the 2023 projected number of AGYW [25].

### Experimental design for a DCE

The attributes of HIVST service were derived from the MoH consolidated guidelines for prevention and treatment of HIV (2022)[5] to conform with the existing DSD models that are currently being implemented for HIVST. However, the distribution model of the HIVST services delivery through the private sector was not considered. Therefore, only four attributes of the HIVST service were considered, of which all had two levels. These include; distribution model (Facility-based versus community-based), delivery channel (primary delivery source versus secondary delivery source), sample/kit type (oral swab versus blood), and level of assistance (assisted versus unassisted) (Table 1). With the four attributes of the HIVST services considered, each with two levels, a full factorial experimental design would give rise to 16 choice sets (2^4^). However, only eight choice sets (fractional factorial) were considered as it has been reported to be optimal to elicit the desired utility measures while preventing cognitive overload of study participants [26,27]. To select the choice sets to be considered, the D-efficient design in STATA version 17.0 (Texas USA) was used and the assumption that no prior coefficient estimates of the attributes in the target population exist (no priors’ matrix) including the alternative specific constant). Given attribute level matrix of 2, 2, 2, 2 (each has two levels) and two alternatives plus the opt-out (opt-out matrix of 1, 1, 1, 1), the modified Fedorov algorithm [28–30], which maximizes the D-efficiency of the design based on covariance matrix of a conditional logit model, was used to generate eight choice sets. The generated choice sets were used to design the DCE questionnaire.

**Table 1:**
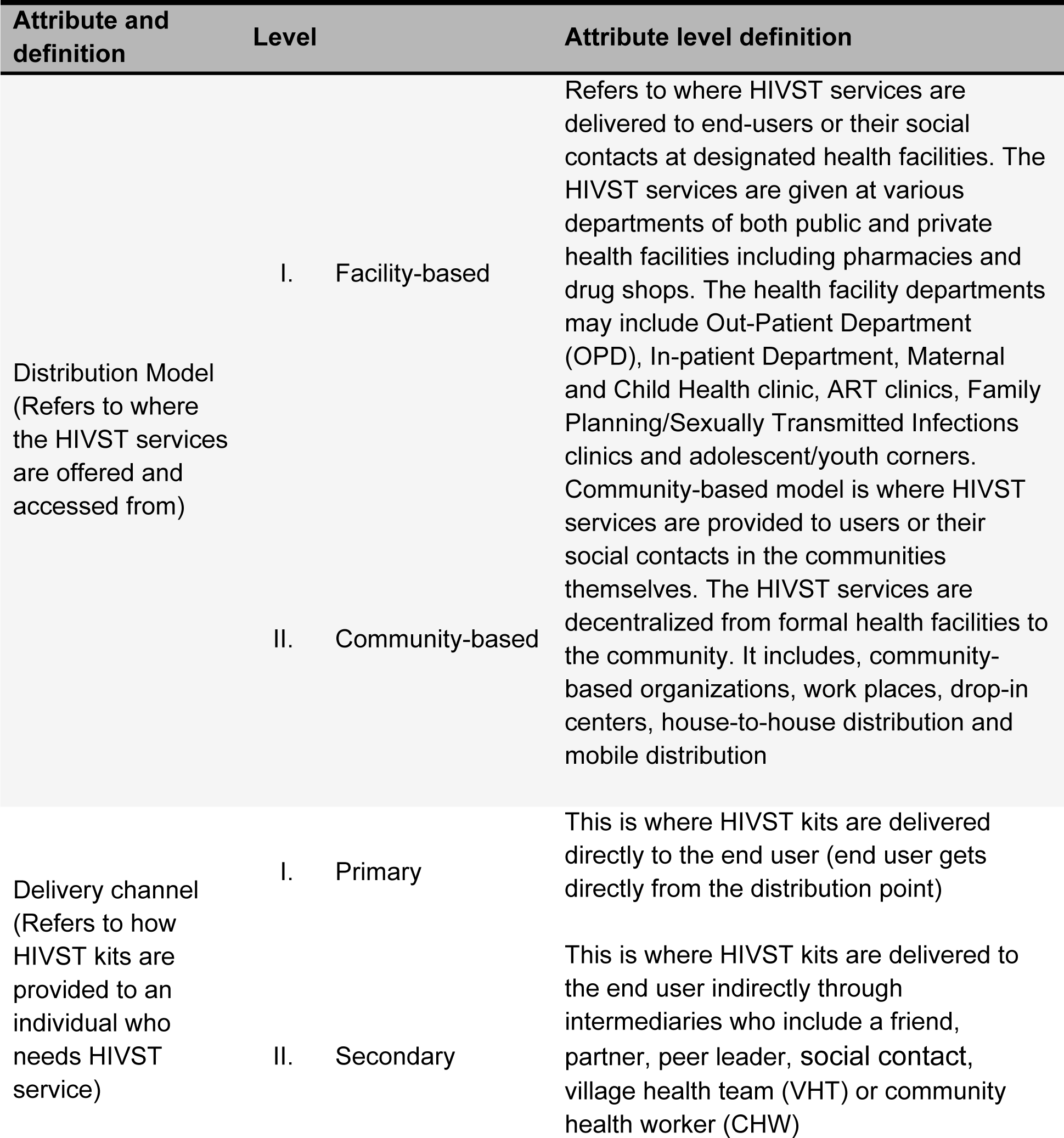
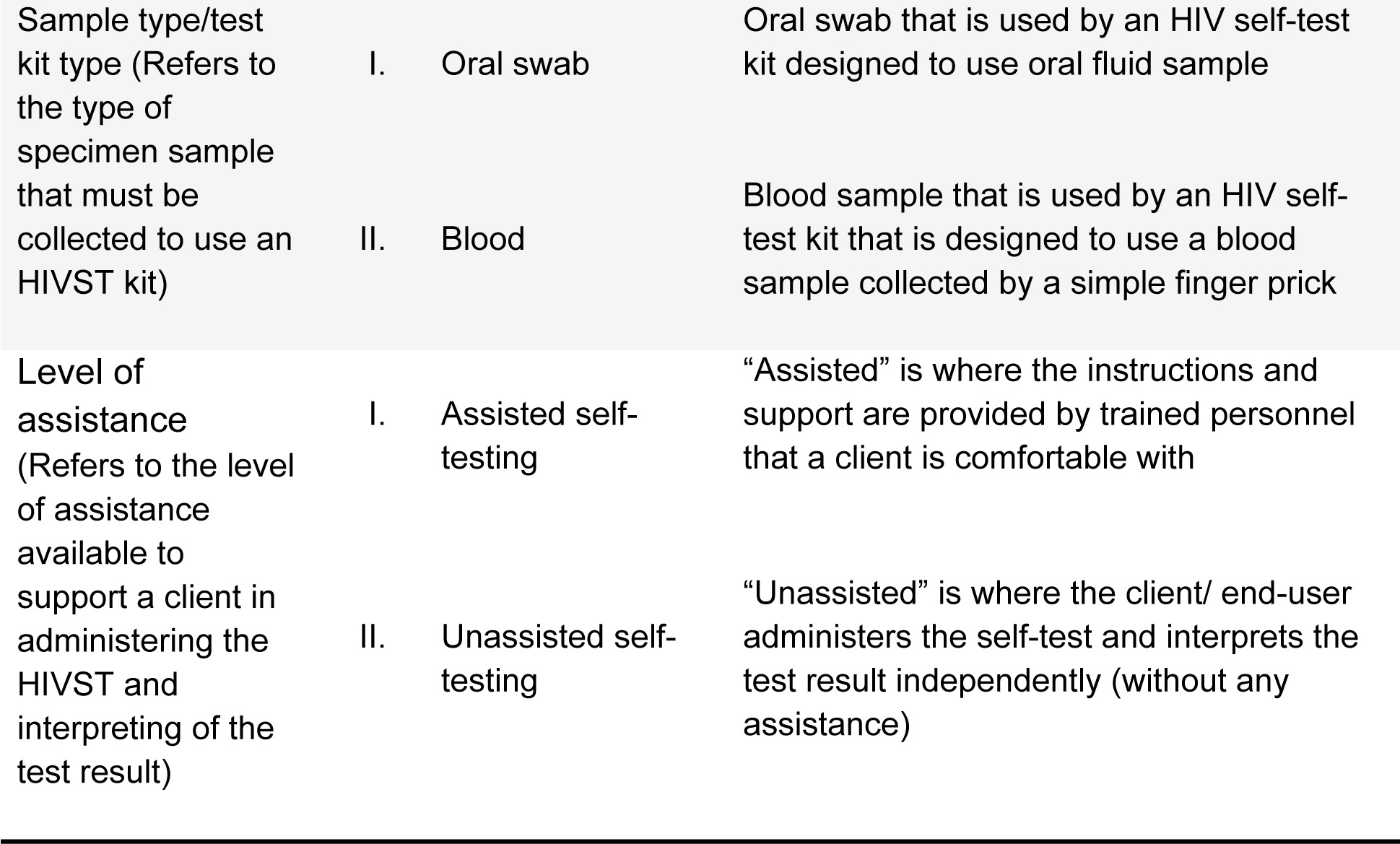
HIVST service attributes and levels adapted from the Consolidated guidelines for prevention and treatment of HIV in Uganda.

### Data collection tool and procedure

A structured questionnaire was used to collect the data on the socio-demographic, and behavioral characteristics, as well as the discrete choices that describe the DSD models for HIVST service. The choice sets of the HIVST service attributes and levels assessed in this study are attached as supplementary file1. The DCE questionnaire was further enhanced with images describing the attributes of HIVST services which were obtained from a public database of images (noun project) [31]. The questionnaire was pretested among 20 HIV high-risk AGYW in Kawempe division in Kampala city, and these participants were excluded in the analysis. A “think-aloud” approach was used in the pretest to assess the participants’ understanding and thought process as they made choices about the tasks presented.

Targeted mobilization was carried out in the study areas to identify AGYW who are at an elevated risk of acquiring HIV. The information about HIVST was given and practical demonstrations of how the HIVST kits are used were done before screening the study participants. Both the oral (OraQuick, OraSure Technologies, Inc. Bethlehem, PA, USA) and blood-based (GOLDEN TIME, Gaobeidian PRISES Biotechnology Co., Ltd, Hebei Province, China) HIVST kits were used in the demonstrations. Trained research assistants administered the questionnaire to AGYW who were eligible for the study.

### Study outcomes

The primary outcome was the choice of preferred alternative of the tasks that offered the highest utility (benefit) to individual participants. The marginal utility gained from each of the attributes was computed as the primary measure. The Relative Importance (RI) was defined as the measure of influence each attribute had on participant’s decision to choose an alternative relative to other attributes in the model. Furthermore, the attribute tradeoffs – defined as the utility of one attribute level a participant is willing to give up (or accept less of) in order to gain a more preferred level of another attribute while still choosing the same alternative was also reported.

### Statistical analysis

The data analysis was done in STATA version 17.0 (Texas, USA). The participant characteristics were summarized using descriptive statistics. For continuous variables, the mean and the standard deviation (SD) or the median and interquartile range (IQR). The categorical variables were summarized using the frequencies and percentages.

The mixed logit model was used to fit a model with the outcome of choice, and attributes of HIVST services as the alternative-specific variables. The relative importance of an HIVST service attribute was computed as a proportion of the range of coefficients of an attribute out of the total sum of the ranges of coefficients of other attributes in the final model [32].

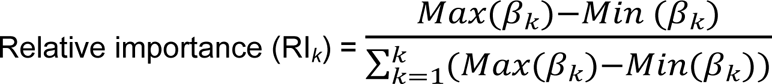

The Trade-offs between one attribute levels to another were computed as a ratio of coefficient of the preferred level to the sacrificed level [33].

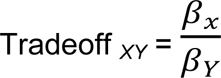

#### Ethical considerations

The study was approved by the Makerere University School of Medicine Research Ethics Committee (Reference: Mak-SOMREC-2023-843), and further clearance was obtained from the Uganda National Council for Science and Technology (UNCST) (Reference: HS5167ES). Administrative approvals to conduct the study were secured from the Kampala Capital City Authority (KCCA) and the Municipal Health Offices of Entebbe and Nansana in Wakiso District. Written informed consent was obtained from all adult participants and emancipated minors. For other participants below 18 years, written informed assent was obtained alongside written informed consent from their guardians, in accordance with UNCST guidelines [34].

## Results

### Socio-demographic characteristics of study participants

The mean age of the participants was 19.8 years (SD: 2.6), about two-thirds (65.9%) were out of school and 71.8 didn’t live with their primary partner. The participants had a median monthly income of about United Stated Dollars $42.1 (IQR: 28.1, 56.1). Slightly more than half of participants (56.8%) had ever been pregnant and about a similar proportion (55.9%) were using a modern contraceptive method. Half of the participants (50.5%) had primary partners who were older by >5 years. 18.8% reported to have experienced intimate partner violence with 10.6% experiencing sexual violence from their intimate partners in the past six months (Table 2).

**Table 2:** Socio-demographic characteristics of AGYW who participated in the study.

| Variable | Category | Frequency<br>(N=340) | Percentage %) |
| --- | --- | --- | --- |
| Age (Years) | Mean (SD) | 19.8 (2.6) |  |
| Schooling status | In school | 116 | 34.1 |
|  | Not in school | 224 | 65.9 |
| Live with partner | Yes | 96 | 28.2 |
|  | No | 244 | 71.8 |
| Monthly income* | Median (IQR) | \$42.1 (28.1, 56.1) | |
| Ever pregnant | Yes | 193 | 56.8 |
|  | No | 147 | 43.2 |
| Contraceptive use<br>in past six months | Yes | 190 | 55.9 |
|  | No | 150 | 44.1 |
| Age difference with<br>primary partner | ≤5 Years | 142 | 41.8 |
|  | >5 Years | 198 | 58.2 |
| Experienced intimate partner violence in past six months | Yes | 64 | 18.8 |
|  | No | 276 | 81.2 |
| Experienced sexual violence in past six months | Yes | 36 | 10.6 |
|  | No | 304 | 89.4 |

\*USD 0.00028 = 1 Uganda shillings

### Behavioral characteristics of the study participants

In the six months preceding the study, 55.6% of participants reported a sexually transmitted infection, 30.3% had multiple sexual partners, and 14.1% reported consistent condom use. Transactional sex was reported by 29.7%, anal sex by 11.2%, and intravenous drug use by 15.6%. Only 6.2% and 4.7% reported using PEP and PrEP, respectively. Overall, 83.2% had ever tested for HIV, and 75.6% of these had done so within the past 12 months. While 71.2% had heard of HIV self-testing (HIVST), only 50.4% had ever used an HIVST kit, and 58.8% reported being unable to obtain one when needed. (Table 3)

**Table 3:**
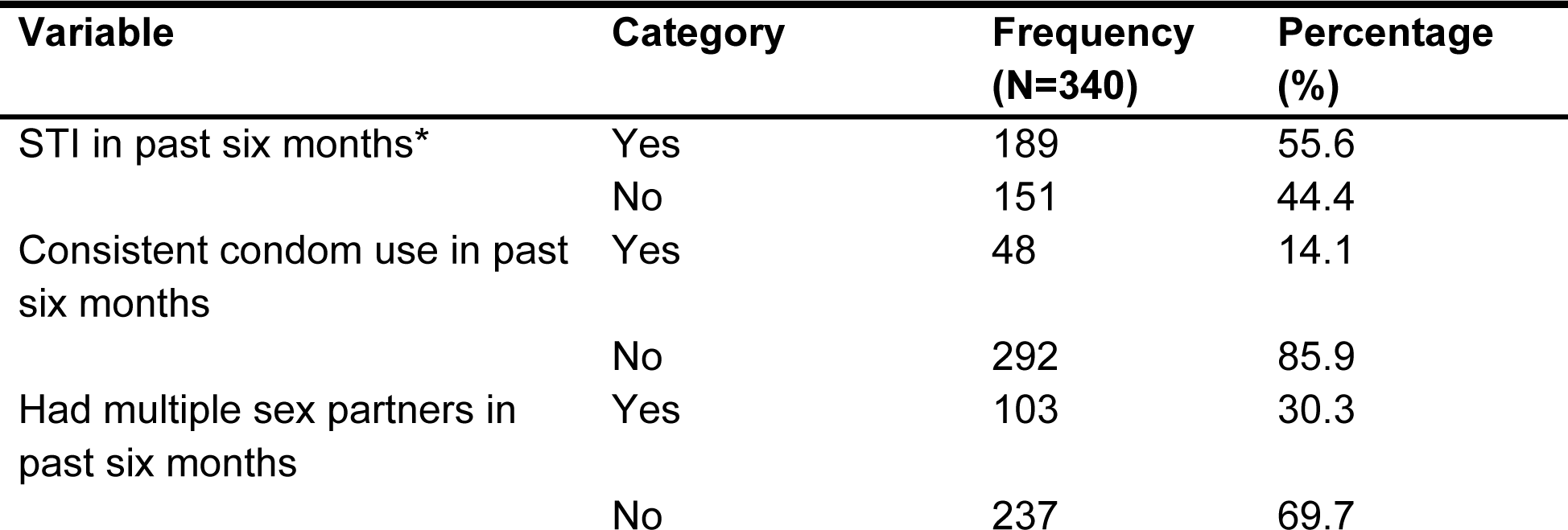
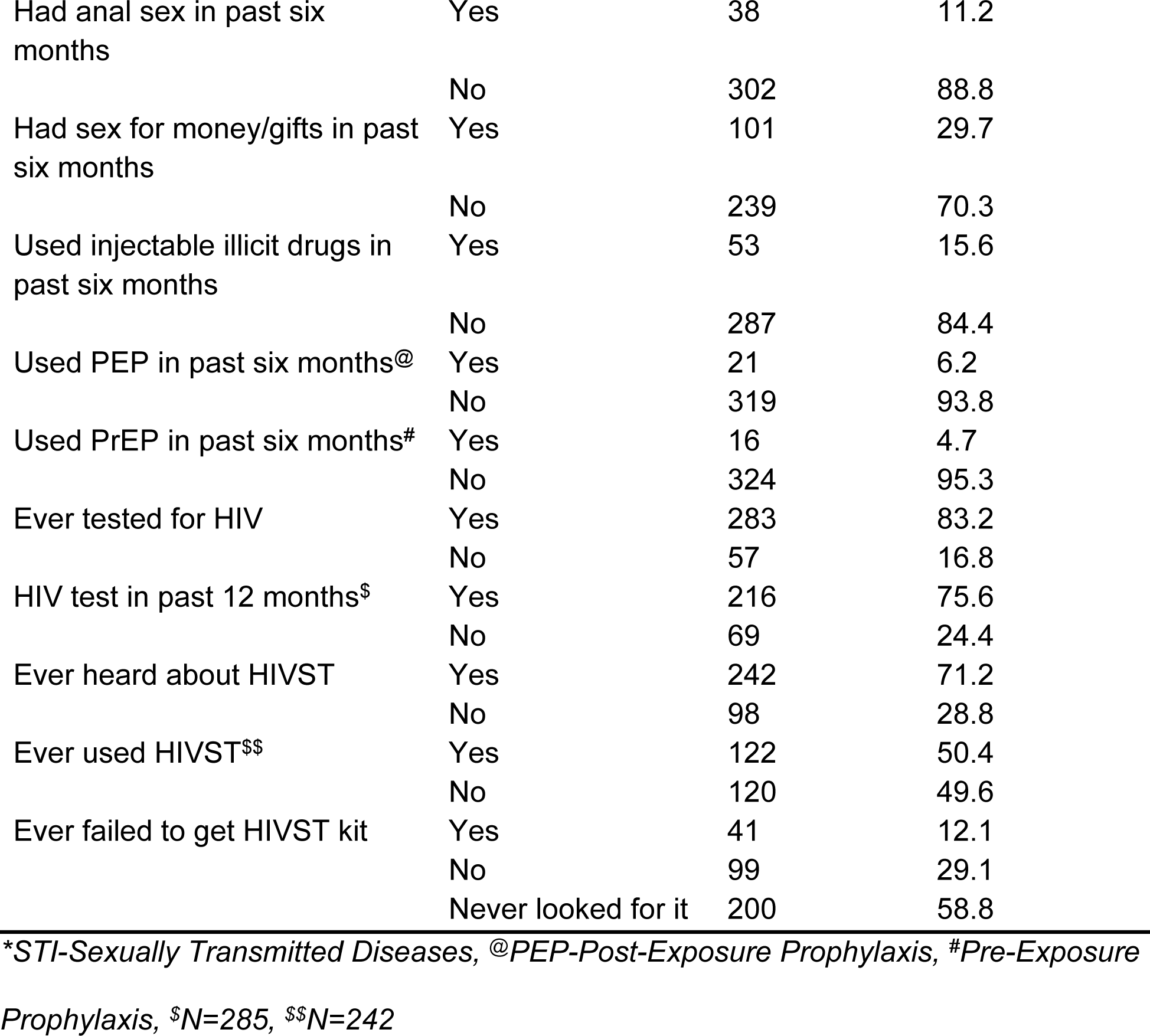
Behavioral characteristics of AGYW who participated in the study.

| Variable | Category | Frequency (N=340) | Percentage (%) |
| --- | --- | --- | --- |
| STI in past six months* | Yes | 189 | 55.6 |
|  | No | 151 | 44.4 |
| Consistent condom use in past six months | Yes | 48 | 14.1 |
|  | No | 292 | 85.9 |
| Had multiple sex partners in past six months | Yes | 103 | 30.3 |
|  | No | 237 | 69.7 |
| Had anal sex in past six months | Yes | 38 | 11.2 |
|  | No | 302 | 88.8 |
| Had sex for money/gifts in past six months | Yes | 101 | 29.7 |
|  | No | 239 | 70.3 |
| Used injectable illicit drugs in past six months | Yes | 53 | 15.6 |
|  | No | 287 | 84.4 |
| Used PEP in past six months@ | Yes | 21 | 6.2 |
|  | No | 319 | 93.8 |
| Used PrEP in past six months# | Yes | 16 | 4.7 |
|  | No | 324 | 95.3 |
| Ever tested for HIV | Yes | 283 | 83.2 |
|  | No | 57 | 16.8 |
| HIV test in past 12 months\$ | Yes | 216 | 75.6 |
|  | No | 69 | 24.4 |
| Ever heard about HIVST | Yes | 242 | 71.2 |
|  | No | 98 | 28.8 |
| Ever used HIVST\$\$ | Yes | 122 | 50.4 |
|  | No | 120 | 49.6 |
| Ever failed to get HIVST kit | Yes | 41 | 12.1 |
|  | No | 99 | 29.1 |
|  | Never looked for it | 200 | 58.8 |

\*STI-Sexually Transmitted Diseases, @PEP-Post-Exposure Prophylaxis, #Pre-Exposure Prophylaxis, \$N=285, \$\$N=242

### Preference of the HIVST service attributes

Participants preferred HIVST services with a facility-based distribution model (coefficient: 0.14; [95% CI: 0.06 - 0.23]), primary delivery (coefficient: 0.09; [95% CI: 0.00 - 0.17]), and unassisted testing (coefficient: 0.46; [95% CI: 0.37 - 0.54]). The type of test kit/sample did not significantly influence participants’ choices.

The level of assistance was the most important attribute that influenced the choice of an alternative (RI: 62.2%), followed by distribution model (RI: 18.9%), delivery channel (RI: 12.2%) and sample/test kit type. (Table 4)

**Table 4:**
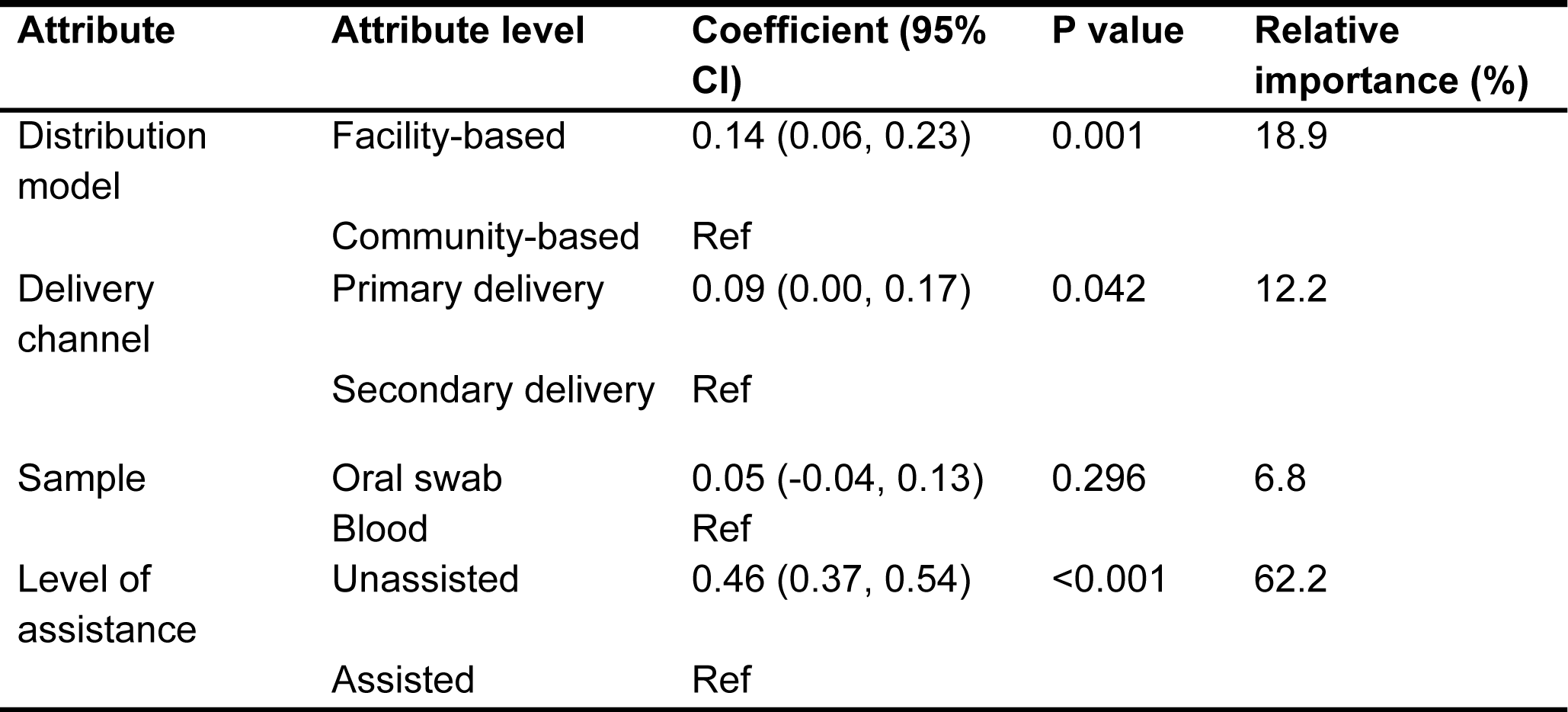
HIV self-testing service preferences of 340 AGYW who participated in the study.

| Attribute | Attribute level | Coefficient (95% CI) | P value | Relative importance (%) |
| --- | --- | --- | --- | --- |
| Distribution model | Facility-based | 0.14 (0.06, 0.23) | 0.001 | 18.9 |
|  | Community-based | Ref |  |  |
| Delivery channel | Primary delivery | 0.09 (0.00, 0.17) | 0.042 | 12.2 |
|  | Secondary delivery | Ref |  |  |
| Sample | Oral swab | 0.05 (-0.04, 0.13) | 0.296 | 6.8 |
|  | Blood | Ref |  |  |
| Level of assistance | Unassisted | 0.46 (0.37, 0.54) | <0.001 | 62.2 |
|  | Assisted | Ref |  |  |

### Willingness to trade-off for a more preferred HIVST attribute

Participants were willing to sacrifice 9.2 units of utility for sample type, 5.1 units for delivery channel, and 3.4 units for distribution model in order to access unassisted HIVST, without any loss in overall utility. However, they were not willing to sacrifice any HIVST attributes in order to obtain a preferred test kit (sample type) (Figure 1).

**Figure 1:**
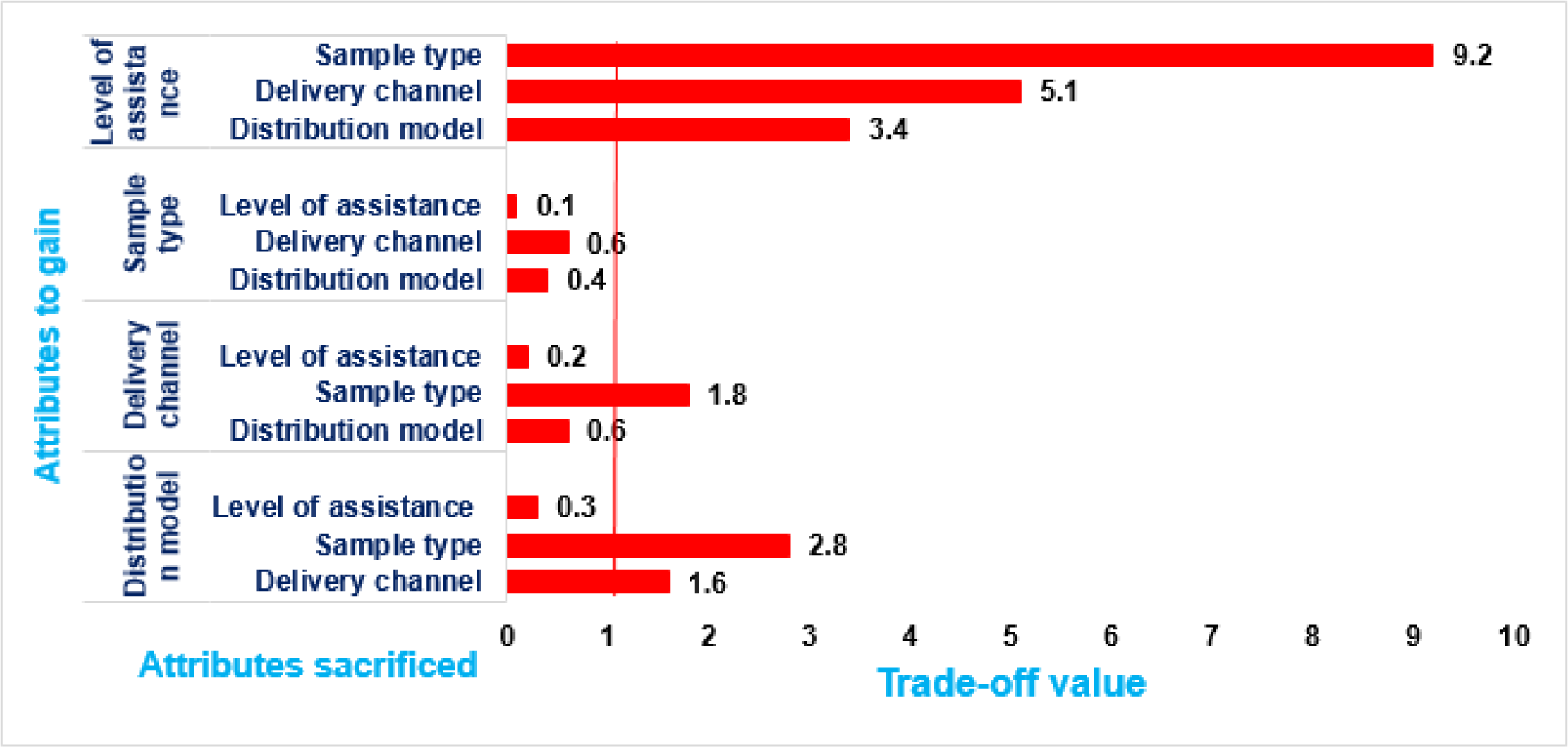
HIV self-testing attribute trade-offs for AGYW.

## Discussion

This DCE explored HIVST preferences among high-risk AGYW in urban Uganda to identify optimal service delivery models. Findings show a strong preference for unassisted HIVST, facility-based distribution, and primary-level delivery channels, while the type of test kit (oral vs. blood) had no significant influence on choice of the alternative tasks. These results provide actionable evidence to guide HIVST models that are responsive to the needs and realities of AGYW.

The level of assistance emerged as the most influential attribute, contributing over 60% of the relative importance in choice decisions. For HIV high-risk AGYW, the ability to self-test without assistance was by far the most important factor in deciding whether to use HIVST service model. It was three times more important than the source of the kit, five times more important than the delivery channel, and nine times more important than the type of test kit. This indicates that even if kit type or distribution method is not ideal, AGYW are still more likely to use HIVST if they can do it without assistance. This finding aligns with global evidence that autonomy and privacy are key facilitators of HIVST uptake [35,36]. Unassisted testing offers control over the testing process while minimizing stigma from health workers, peers, or partners – barriers to which AGYW are particularly vulnerable [37,38]. It also reduces practical obstacles such as scheduling, travel, and waiting times, while allowing immediate testing at moments of perceived risk. HIVST models targeting AGYW should therefore, prioritize unassisted testing to maximize acceptability and uptake.

The distribution model provided participants with significant utility and ranked second in relative importance. While unassisted testing was the dominant driver of choice, the location from which test kits are obtained also played a key role in decision-making. Health facilities are often viewed as reliable sources of high-quality HIVST kits, with studies in SSA reporting strong trust in facility-based quality assurance [39–41], even when individuals prefer to test privately [41]. In addition, some young women may perceive facility staff as bound by confidentiality rules, which helps reduce fears of unwanted disclosure. Integrating unassisted HIVST access within trusted health facilities – particularly at primary care level – may therefore, maximize uptake. Furthermore, the efforts to further strengthen trust in these facilities among AGYW could enhance both HIVST use and broader service engagement.

In this study, delivering HIVST kits through primary delivery channels provided greater utility than secondary distribution. AGYW preferred to collect test kits themselves rather than relying on others, partly because direct pick-up minimizes risks of unintended disclosure, reduces anxiety, and allows them autonomy to decide when to collect and use the HIVST kit. Evidence from Zimbabwe and Malawi similarly shows that youth prefer collecting kits from health facilities or pharmacies because of trust, confidentiality, and independence. While secondary distribution – particularly partner-delivered kits – has proven effective [42][43], it may be less suitable for unmarried youth or those facing high stigma, as it can compromise privacy. Importantly, the willingness of AGYW to pick up HIVST kits directly from trusted facilities creates opportunities to integrate kit distribution with other health services such as sexual and reproductive health counselling, family planning, STI clinics, and HIV prevention platforms including PrEP and PEP delivery – strengthening both reach and utility of other prevention commodities.

The sample type (oral versus blood-based kits) was not a significant driver of choice and carried the lowest relative importance. For AGYW, the biological sample collection method was less influential compared to other features such as level of assistance, distribution model, and delivery channel. Similar to our findings, other studies have also ranked sample type lower than factors such as cost, access point, and level of assistance [44]. While some contexts suggest that oral swabs may be more acceptable due to their non-invasive nature [45,46], our results indicate that among AGYW, other HIVST features outweigh the sample collection method. Although blood-based kits are sometimes perceived as more accurate [47], oral kits are often viewed as easier and less painful. In Uganda, both types are approved for use, though oral HIVST kits remain more expensive. This creates an opportunity for implementers to expand access by making affordable blood-based HIVST kits available to young women, thereby supporting broader HIV testing uptake.

## Limitations

This study employed a DCE to elicit stated preferences using hypothetical scenarios, which may not fully reflect actual behavior when AGYW are offered HIVST options. This could result in either over- or under-estimation of preferences. Furthermore, as HIVST becomes normalized, AGYW’s preferences may evolve over time; however, the cross-sectional design of this study did not allow assessment of such temporal trends. Although the attributes and levels were guided by MoH-recommended DSD models, the potential influence of cost, an important determinant of HIVST uptake, was not assessed. Some data were collected retrospectively and are subject to recall bias, though this was minimized by restricting the recall period to six months (except for HIV testing history, which extended to 12 months). Consecutive sampling, a non-probability technique, may have introduced selection bias by not giving every eligible participant an equal chance of inclusion. Similarly, the exclusion of participants who did not speak English or Luganda could have introduced bias; however, Luganda is widely spoken in the study setting. Finally, the study focused on HIV high-risk AGYW in urban settings, which may limit the generalizability of findings to rural or lower-risk populations. Despite these limitations, the potential biases are unlikely to have substantially affected the validity of the findings.

## Conclusions

HIV high-risk AGYW living in urban settings showed a clear preference for unassisted HIVST, delivered through a facility-based distribution model and a primary-level delivery channel. While the type of test kit was less influential, the autonomy and convenience afforded by unassisted testing emerged as the strongest driver of HIVST service choice.

## Recommendations

From the findings, we recommend that Ministries of Health and HIV program implementing partners to integrate unassisted HIVST into scale-up strategies for AGYW, ensuring that test kits are packaged with clear, youth-friendly instructions. This should be coupled with strengthening facility-based distribution, particularly through youth-friendly facilities, pharmacies, and drug shops with extended hours, to maximize accessibility. Such approaches will expand opportunities for AGYW to conveniently and confidentially obtain HIVST kits from trusted sources.

Integration of HIVST into existing health-facility platforms – including sexual and reproductive health counselling, family planning, STI clinics, and HIV prevention services such as PrEP delivery sites – can further enhance distribution to HIV high-risk AGYW.

Given that test kit type was not a significant driver of choice, Ministries of Health should focus on ensuring the availability and affordability of multiple kit options, rather than restricting scale-up to a single type.

Future research should explore the influence of additional attributes not measured in this study, such as cost and willingness to pay, which may directly affect uptake.

Moreover, as HIVST becomes increasingly normalized, longitudinal research will be important to monitor how preferences evolve over time.

## Data Availability

The dataset has been submitted as a supporting file

## Acknowledgements

We gratefully acknowledge all the participants who generously gave their time to take part in this study. We also extend our appreciation to the Village Health Teams (VHTs) in Kampala and Wakiso districts for their invaluable support in mobilizing study participants.

## Notes

### Competing Interest Statement

The authors have declared no competing interest.

### Funding Statement

The findings and data reported in this publication were supported by the Fogarty International Center and the National Institute of Mental Health of the National Institutes of Health under Award Number D43 TW010037. The contents are solely the responsibility of the authors and do not necessarily represent the official views of the National Institutes of Health. The funders had no role in the study conceptualization, design, data collection and analysis, decision to publish or preparation of the Manuscript. FCS received the funding for this work.

### Author Declarations

The study was approved by the Makerere University School of Medicine Research Ethics Committee (Reference: Mak-SOMREC-2023-843), and further clearance was obtained from the Uganda National Council for Science and Technology (UNCST) (Reference: HS5167ES). Administrative approvals to conduct the study were secured from the Kampala Capital City Authority (KCCA) and the Municipal Health Offices of Entebbe and Nansana in Wakiso District. Written informed consent was obtained from all adult participants and emancipated minors. For other participants below 18 years, written informed assent was obtained alongside written informed consent from their guardians, in accordance with UNCST guidelines.

## References

1. UNAIDS. Fact Sheet 2025. 2025 [cited 8 Aug 2025]. Available: https://www.unaids.org/sites/default/files/2025-07/2025_Global_HIV_Factsheet_en.pdf

2. Uganda AIDS Commission. 2024 Uganda HIV and AIDS FActsheet. 2024 [cited 8 Aug 2025]. Available: https://uac.go.ug/index.php/easy-customization/custom-404-page-and-offline-page

3. Mwine P, Kwesiga B, Migisha R, Kabwama S, Cheptoris J, Mudiope P, et al. HIV Positivity Rate and Recent HIV Infections Among Adolescent Girls and Young Women 10-24 years, Uganda, 2017-2021. 2022.

4. UNAIDS. Understanding measures of progress towards the 95–95–95 HIV testing, treatment and viral suppression targets. 2023 [cited 8 Aug 2025]. Available: https://www.unaids.org/sites/default/files/media_asset/progress-towards-95-95-95_en.pdf

5. Ministry of Health. Consolidated guidelines for the prevention and treatment of HIV and AIDS in Uganda. In: MoH-Uganda. Kampala; 2022.

6. MoH. Addendum to the HIV testing services policy & implementation guidelines for HIV self-testing & Assisted Partner Notification services. In: Ministry of Health Uganda [Internet]. Kampala; 2018 [cited 13 May 2023]. Available: https://www.health.go.ug/download-attachment/Sv3MJWLFryzTR69NvhNTzDuglek9ELnr5vfct1_pwYE

7. Murewanhema G, Musuka G, Moyo P, Moyo E, Dzinamarira T. HIV and adolescent girls and young women in sub-Saharan Africa: A call for expedited action to reduce new infections. IJID Reg. 2022;5: 30–32. doi:10.1016/j.ijregi.2022.08.009

8. Kelvin EA, Akasreku B. The evidence for HIV self-testing to increase HIV testing rates and the implementation challenges that remain. Curr HIV/AIDS Rep. 2020;17: 281–289.

9. UNAIDS. HIV self-testing: what you need to know. 2024 [cited 8 Aug 2025]. Available: https://www.unaids.org/sites/default/files/hiv-self-testing-what-you-need-to-know_en.pdf

10. Fischer AE, Abrahams M, Shankland L, Lalla-Edward ST, Edward VA, De Wit J. The evolution of HIV self-testing and the introduction of digital interventions to improve HIV self-testing. Front Reprod Heal. 2023;5: 1121478. doi:10.3389/frph.2023.1121478

11. World Health Organization. HIV Self-testing and Partner Notification. Supplement to Consolidated guidelines on HIV testing services. In: Geneva, Switzerland [Internet]. 2016 [cited 18 May 2025]. Available: https://apps.who.int/iris/bitstream/handle/10665/251655/9789241549868-eng.pdf;jsessionid=A4502F66DB65552D95D583F71AC7468D?sequence=1

12. Jamil MS, Eshun-Wilson I, Witzel TC, Siegfried N, Figueroa C, Chitembo L, et al. Examining the effects of HIV self-testing compared to standard HIV testing services in the general population: a systematic review and meta-analysis. EClinicalMedicine. 2021;38.

13. Witzel TC, Eshun-Wilson I, Jamil MS, Tilouche N, Figueroa C, Johnson CC, et al. Comparing the effects of HIV self-testing to standard HIV testing for key populations: a systematic review and meta-analysis. BMC Med. 2020;18: 381. doi:10.1186/s12916-020-01835-z

14. McGee K, d’Elbée M, Dekova R, Sande LA, Dube L, Masuku S, et al. Costs of distributing HIV self-testing kits in Eswatini through community and workplace models. BMC Infect Dis. 2024;22: 976. doi:10.1186/s12879-023-08694-y

15. Matsimela K, Sande LA, Mostert C, Majam M, Phiri J, Zishiri V, et al. The cost and intermediary cost-effectiveness of oral HIV self-test kit distribution across 11 distribution models in South Africa. BMJ Glob Heal. 2021;6. doi:10.1136/bmjgh-2021-005019

16. Lancaster KJ. A New Approach to Consumer Theory Journal of Political Economy, 74 (2). S; 1966.

17. McFadden D. Conditional logit analysis of qualitative choice behavior. 1972.

18. Abiiro GA, Leppert G, Mbera GB, Robyn PJ, De Allegri M. Developing attributes and attribute-levels for a discrete choice experiment on micro health insurance in rural Malawi. BMC Health Serv Res. 2014;14: 235. doi:10.1186/1472-6963-14-235

19. Szinay D, Cameron R, Naughton F, Whitty JA, Brown J, Jones A. Understanding Uptake of Digital Health Products: Methodology Tutorial for a Discrete Choice Experiment Using the Bayesian Efficient Design. J Med Internet Res. 2021;23: e32365. doi:10.2196/32365

20. Maulide Cane R, Melesse DY, Kayeyi N, Manu A, Wado YD, Barros A, et al. HIV trends and disparities by gender and urban–rural residence among adolescents in sub-Saharan Africa. Reprod Health. 2021;18: 120. doi:10.1186/s12978-021-01118-7

21. UBOS. National Population and Housing Census 2024, Final Report. Kampala; 2024.

22. Mukwaya PI, Mbabazi J, Ernstson H. Kampala: city report. Kampala; 2025.

23. Sekhon M, Cartwright M, Francis JJ. Acceptability of healthcare interventions: an overview of reviews and development of a theoretical framework. BMC Health Serv Res. 2017;17: 1–13.

24. Orme B. Sample size issues for conjoint analysis studies. Sequim Sawtooth Softw Tech Pap. 1998.

25. Uganda Breau of Statistics (UBOS). Gender statistics portal; one-stop center for gender statistics in Uganda. 2023 [cited 24 Nov 2023]. Available: http://gender.ubos.org:8080/

26. Wong SF, Norman R, Dunning TL, Ashley DM, Lorgelly PK. A protocol for a discrete choice experiment: understanding preferences of patients with cancer towards their cancer care across metropolitan and rural regions in Australia. BMJ Open. 2014;4: e006661.

27. Makabayi-Mugabe R, Musaazi J, Zawedde-Muyanja S, Kizito E, Namwanje H, Aleu P, et al. Developing a patient-centered community-based model for management of multi-drug resistant tuberculosis in Uganda: a discrete choice experiment. BMC Health Serv Res. 2022;22: 154. doi:10.1186/s12913-021-07365-5

28. Carlsson F, Martinsson P. Design techniques for stated preference methods in health economics. Health Econ. 2003;12: 281–294.

29. Cook RD, Nachtrheim CJ. A comparison of algorithms for constructing exact D-optimal designs. Technometrics. 1980;22: 315–324.

30. Zwerina K, Huber J, Kuhfeld WF. A general method for constructing efficient choice designs. Durham, NC Fuqua Sch Business, Duke Univ. 1996;7.

31. Noun project. Photos for everything. 2022 [cited 3 Sep 2023]. Available: https://thenounproject.com/

32. Orme B. Getting started with conjoint analysis: strategies for product design and pricing research second edition. Madison Res Publ LLC. 2010.

33. Hensher DA, Rose JM, Greene WH. Applied choice analysis. Cambridge university press; 2015.

34. UNCST. National Guidelines for Research involving Humans as Research Participants. Uganda: Uganda National Council of Science and Technology; 2014 [cited 22 Nov 2023]. Available: https://www.uncst.go.ug/manage/files/downloads/Human Subjects Protection Guidelines July 2014(1).pdf

35. Njau B, Covin C, Lisasi E, Damian D, Mushi D, Boulle A, et al. A systematic review of qualitative evidence on factors enabling and deterring uptake of HIV self-testing in Africa. BMC Public Health. 2019;19: 1289. doi:10.1186/s12889-019-7685-1

36. Mukora-Mutseyekwa F, Mundagowa PT, Kangwende RA, Murapa T, Tirivavi M, Mukuwapasi W, et al. Implementation of a campus-based and peer-delivered HIV self-testing intervention to improve the uptake of HIV testing services among university students in Zimbabwe: the SAYS initiative. BMC Health Serv Res. 2022;22: 222. doi:10.1186/s12913-022-07622-1

37. Nyblade L, Ndirangu JW, Speizer IS, Browne FA, Bonner CP, Minnis A, et al. Stigma in the health clinic and implications for PrEP access and use by adolescent girls and young women: conflicting perspectives in South Africa. BMC Public Health. 2022;22: 1916. doi:10.1186/s12889-022-14236-z

38. Jani N, Mathur S, Kahabuka C, Makyao N, Pilgrim N. Relationship dynamics and anticipated stigma: Key considerations for PrEP use among Tanzanian adolescent girls and young women and male partners. PLoS One. 2021;16: e0246717. doi:10.1371/journal.pone.0246717

39. Mugambi ML, Odhiambo BO, Dollah A, Marwa MM, Nyakina J, Kinuthia J, et al. Women’s preferences for HIV prevention service delivery in pharmacies during pregnancy in Western Kenya: a discrete choice experiment. J Int AIDS Soc. 2024;27: e26301.

40. Hatzold K, Gudukeya S, Mutseta MN, Chilongosi R, Nalubamba M, Nkhoma C, et al. HIV self-testing: breaking the barriers to uptake of testing among men and adolescents in sub-Saharan Africa, experiences from STAR demonstration projects in Malawi, Zambia and Zimbabwe. J Int AIDS Soc. 2019;22 Suppl 1: e25244. doi:10.1002/jia2.25244

41. Asiimwe S, Oloya J, Song X, Whalen CC. Accuracy of Un-supervised Versus Provider-Supervised Self-administered HIV Testing in Uganda: A Randomized Implementation Trial. AIDS Behav. 2014;18: 2477–2484. doi:10.1007/s10461-014-0765-4

42. Thirumurthy H, Masters SH, Mavedzenge SN, Maman S, Omanga E, Agot K. Promoting male partner HIV testing and safer sexual decision making through secondary distribution of self-tests by HIV-negative female sex workers and women receiving antenatal and post-partum care in Kenya: a cohort study. lancet HIV. 2016;3: e266–e274.

43. Masters SH, Agot K, Obonyo B, Napierala Mavedzenge S, Maman S, Thirumurthy H. Promoting Partner Testing and Couples Testing through Secondary Distribution of HIV Self-Tests: A Randomized Clinical Trial. PLoS Med. 2016;13: e1002166. doi:10.1371/journal.pmed.1002166

44. Chetty-Makkan CM, Hoffmann CJ, Charalambous S, Botha C, Ntshuntshe S, Nkosi N, et al. Youth Preferences for HIV Testing in South Africa: Findings from the Youth Action for Health (YA4H) Study Using a Discrete Choice Experiment. AIDS Behav. 2021;25: 182–190. doi:10.1007/s10461-020-02960-9

45. Peck RB, Lim JM, Van Rooyen H, Mukoma W, Chepuka L, Bansil P, et al. What should the ideal HIV self-test look like? A usability study of test prototypes in unsupervised HIV self-testing in Kenya, Malawi, and South Africa. AIDS Behav. 2014;18: 422–432.

46. Kurth AE, Cleland CM, Chhun N, Sidle JE, Were E, Naanyu V, et al. Accuracy and Acceptability of Oral Fluid HIV Self-Testing in a General Adult Population in Kenya. AIDS Behav. 2016;20: 870–879. doi:10.1007/s10461-015-1213-9

47. Arije O, Madan J, Hlungwani T. Preferences in adolescents and young people’s sexual and reproductive health services in Nigeria: a discrete choice experiment. Health Econ Rev. 2024;14: 24.

